# The Role of Machine Learning Techniques to Tackle COVID-19 Crisis: A Systematic Review

**DOI:** 10.1101/2020.08.23.20180158

**Authors:** Hafsa Bareen Syeda, Mahanazuddin Syed, Kevin Sexton, Shorabuddin Syed, Salma Begum, Farhan Syed, Feliciano Yu

## Abstract

**Background:** The novel coronavirus responsible for COVID-19 has caused havoc with patients presenting a spectrum of complications forcing the healthcare experts around the globe to explore new technological solutions, and treatment plans. Machine learning (ML) based technologies have played a substantial role in solving complex problems, and several organizations have been swift to adopt and customize them in response to the challenges posed by the COVID-19 pandemic.

**Objective:** The objective of this study is to conduct a systematic literature review on the role of ML as a comprehensive and decisive technology to fight the COVID-19 crisis in the arena of epidemiology, diagnosis, and disease progression.

**Methods:** A systematic search in PubMed, Web of Science, and CINAHL databases was performed according to the Preferred Reporting Items for Systematic Reviews and Meta-analysis (PRISMA) guidelines to identify all potentially relevant studies published and made available between December 1, 2019, and June 27, 2020. The search syntax was built using keywords specific to COVID-19 and ML. A total of 128 qualified articles were reviewed and analyzed based on the study objectives.

**Results:** The 128 publications selected were classified into three themes based on ML applications employed to combat the COVID-19 crisis: Computational Epidemiology (CE), Early Detection and Diagnosis (EDD), and Disease Progression (DP). Of the 128 studies, 70 focused on predicting the outbreak, the impact of containment policies, and potential drug discoveries, which were grouped into the CE theme. For the EDD, we grouped forty studies that applied ML techniques to detect the presence of COVID-19 using the patients’ radiological images or lab results. Eighteen publications that focused on predicting the disease progression, outcomes (recovery and mortality), Length of Stay (LOS), and number of Intensive Care Unit (ICU) days for COVID-19 positive patients were classified under the DP theme.

**Conclusions:** In this systematic review, we assembled the current COVID-19 literature that utilized ML methods to provide insights into the COVID-19 themes, highlighting the important variables, data types, and available COVID-19 resources that can assist in facilitating clinical and translational research.

## Introduction

COVID-19 is a worldwide health crisis, more than 16 million people are infected and caused over 666,000 deaths (up to 29 July 2020) across the globe [1]. The resulting impact on healthcare systems is that many countries have overstretched their resources to mitigate spread of the pandemic [2]. There is an urgent need for effective drugs and vaccines to treat and prevent the infection. Due to the lack of validated therapeutics, most of the containment measures to curtail the spread rely on social distancing, quarantine, and lockdown policies [2–4]. The transmission has been slowed but not eliminated; with ease of restriction, there is a fear of the second wave of infection [5, 6]. To restrict the second potential outbreak, advanced containment measures such as contact tracing, identifying hotspots, etc., are now needed [7, 8].

There is a high degree of variance in the COVID-19 symptoms ranging from mild flu to acute respiratory distress syndrome or fulminant pneumonia [9–11]. Machine Learning (ML) techniques have been employed on different scales ranging from prediction of disease spread trajectory to diagnostic and prognostic models development [12, 13]. A wide range of data types including social media, radiological images, omics data, drug databases, and data collected from public health agencies, etc. have been used for the prediction [1, 14–18]. Several studies focused on reviewing published articles and papers that apply Artificial Intelligence (AI) to fight and support the coronavirus response [12, 13, 19, 20]. One among them is a study by Wynants et al [13] that focused on critical appraisal of models that aimed to predict the risk of developing the disease, hospital admissions, and progression. However, a majority of epidemiological studies that aimed to model disease transmission or fatality rate, etc., were excluded.

The primary aim of this study is to conduct a systematic literature review on the role of ML as a technology to combat the COVID-19 crisis and to assess its application in the epidemiological, clinical, and molecular advancements. Specifically, we summarized the area of application, data types used, types of AI and ML methods employed and their performance, scientific findings, and challenges experienced in adopting this technology.

## Methods

This systematic literature review followed the guidelines of the Preferred Reporting Items for Systematic Reviews and Meta-Analyses (PRISMA) framework for preparation and reporting [21].

### Eligibility Criteria

This study focused on peer-reviewed publications, as well as, preprints that applied ML techniques to analyze and address COVID-19 crisis on different scales including diagnostics, prognostics, disease spread forecast, omics, and drug development.

### Data sources and search strategy

PubMed, Web of Science, and the CINAHL databases were searched, restricting the search to research articles published in English and in peer-reviewed or preprint journals or conference proceedings available from Dec 1, 2019, through June 27, 2020. The search syntax was built with the guidance of a professional librarian and included the following search terms: “CORONAVIRUS”, “COVID-19”, “covid19”, “cov-19”, “cov19”, “severe acute respiratory syndrome coronavirus 2”, “Wuhan coronavirus”, “Wuhan seafood market pneumonia virus”, “coronavirus disease 2019 virus”, “SARS-CoV-2”, “SARS2”, “SARS-2”, “2019-nCoV”, “2019 novel coronavirus”, “novel corona”, “Machine Learning”, “Artificial Intelligence”, “Deep Learning”, and “Neural Network”. Refer to Multimedia Appendix 1 for search query syntax. Figure 1 illustrates the process of identifying eligible publications.

**Figure 1.**
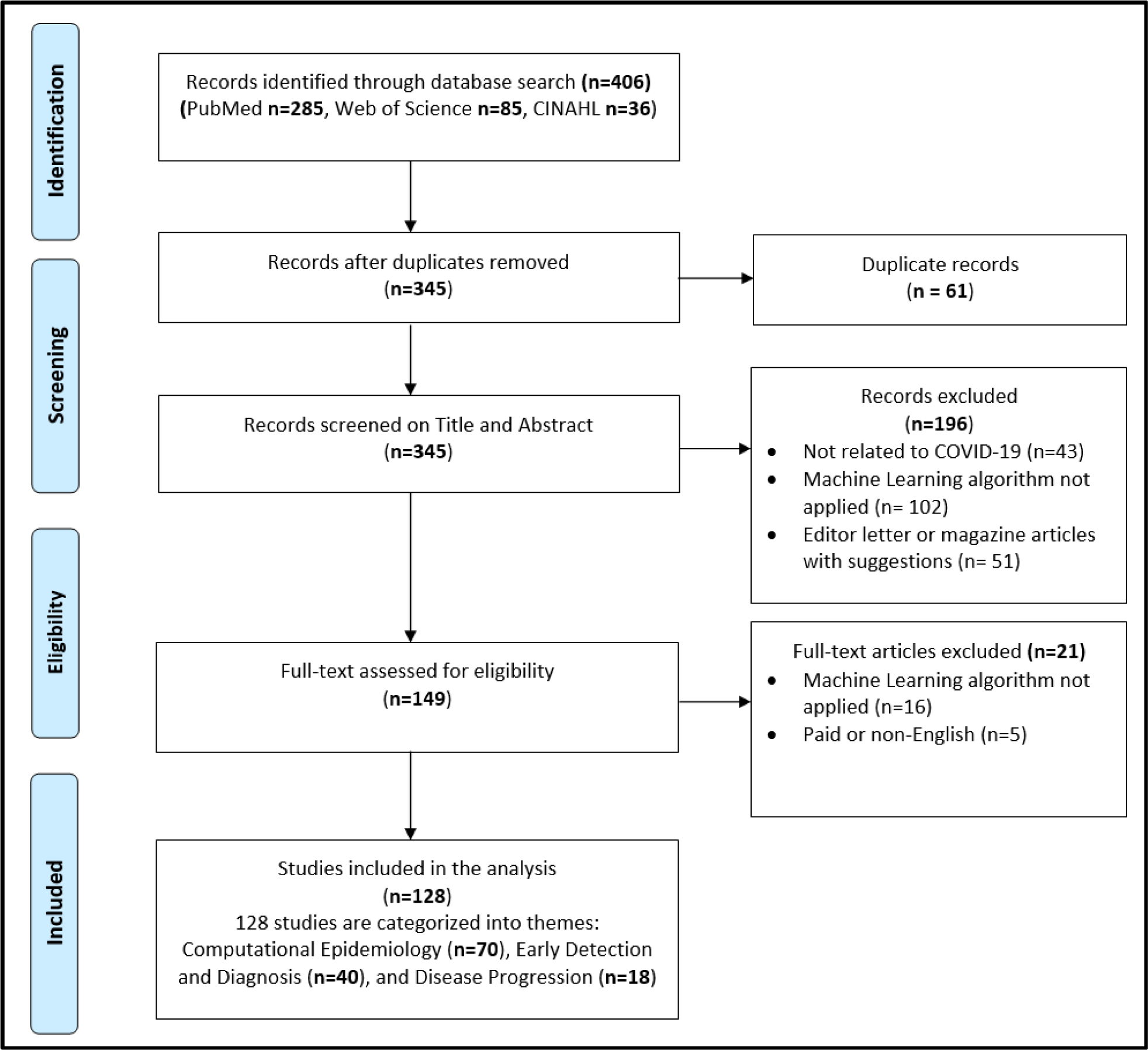
PRISMA flow diagram of systematic identification, screening, eligibility, and inclusion of publications that applied ML techniques to tackle COVID-19 pandemic.

### Study selection

Following the systematic search process, 406 publications were retrieved. Of that, 61 duplicate publications were removed, leaving 345 potentially relevant articles for the title and abstract screening. Two teams (HB, SS and MS, SB) screened these articles independently, leading to the removal of another 196 publications, and 149 publications were retained for a full-text assessment. These were assessed for eligibility, resulting in 128 total publications that were included in the final analysis. Disagreements were resolved by an independent review by third person (FS).

### Data collection and analysis

A qualitative and quantitative descriptive analysis was performed on the qualified studies that had applied ML techniques for tackling COVID-19 pandemic. Based on the area of application, the studies were categorized into themes: 1) Computational Epidemiology (CE), 2) Early Detection and Diagnosis (EDD), and 3) Disease Progression (DP). Qualitative analysis was done on studies from the CE and quantitative descriptive analysis was done for studies that belonged to the EDD and DP themes. After the extraction and analysis, we summarized and reported the findings in tables and figures in accordance with the aim of the study.

## Results

The 128 publications selected were categorized into three themes based on the ML applications employed to combat COVID-19 crisis. The three themes (CE, EDD, and DP) were identified based on ML techniques used to predict, classify, assess, track, and control the spread of the virus. Description of each theme and related publications is presented in Table 1. During the initial days of COVID-19 spread, the majority of the studies focused on predicting the outbreak and potential drug discoveries, which we identified in 70 studies, and classified them into the CE. Forty studies that applied ML techniques to detect COVID-19 using patients’ radiological images or lab results were grouped into EDD. We identified 18 publications that focused on predicting disease progression, outcomes (recovery and mortality), Length of Stay (LOS), and the number of Intensive Care Unit (ICU) days for COVID-19 positive patients, which are grouped under DP theme. Overtime trend of COVID-19 articles by month and themes is shown in Figure 2, which depicts an initial surge of publications focusing on theme CE and then followed by EDD.

**Table 1.**
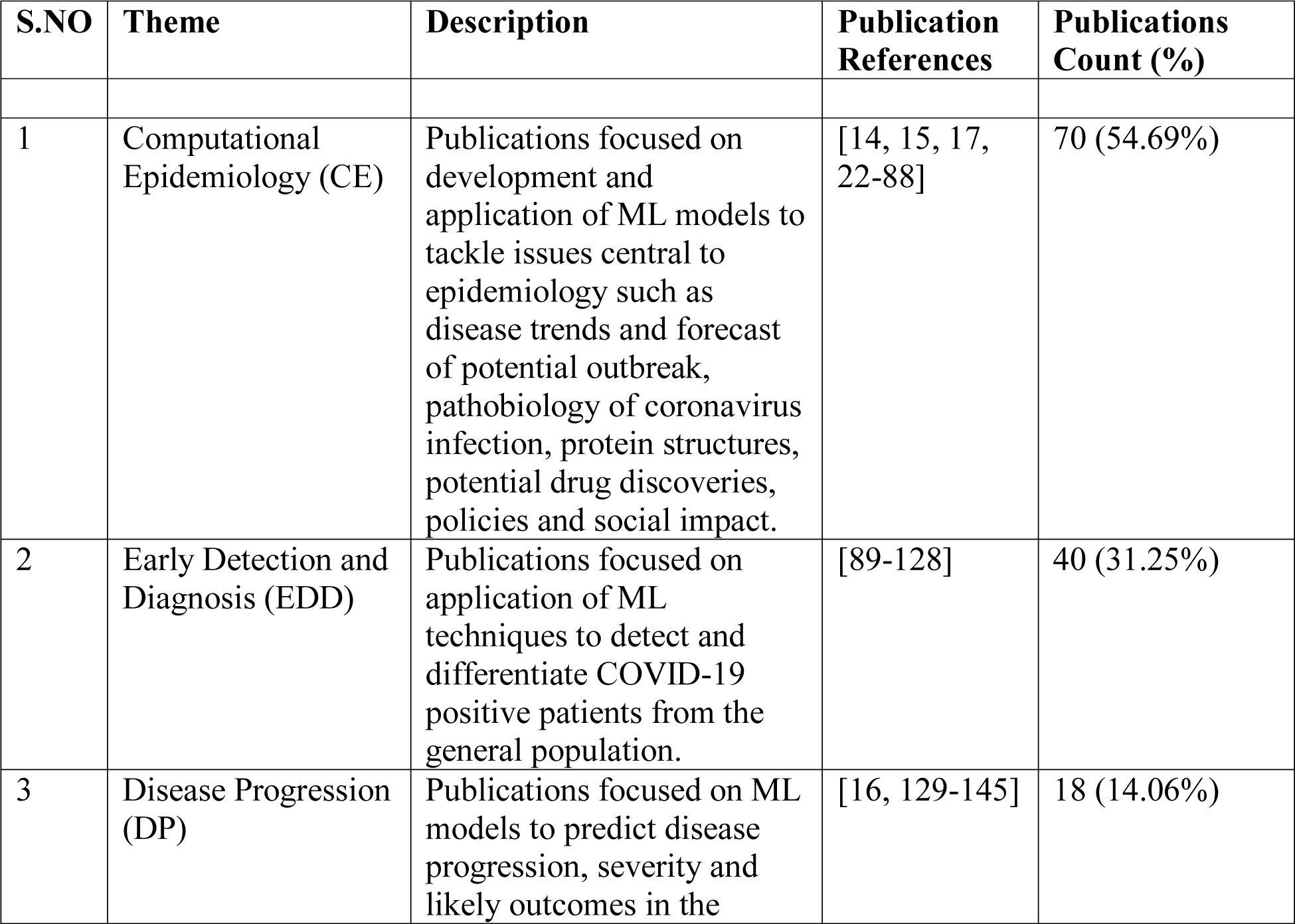
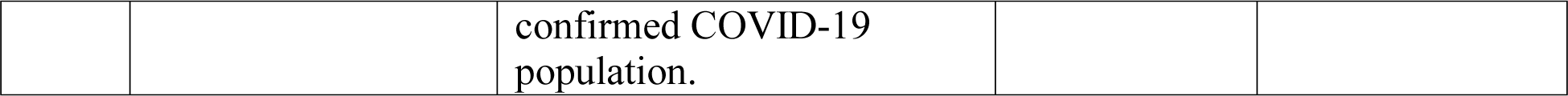
An overview of the 128 papers analyzed in the literature, indicating the three themes and their description. The themes are listed according to the frequency of publication percentage and total count.

**Figure 2.**
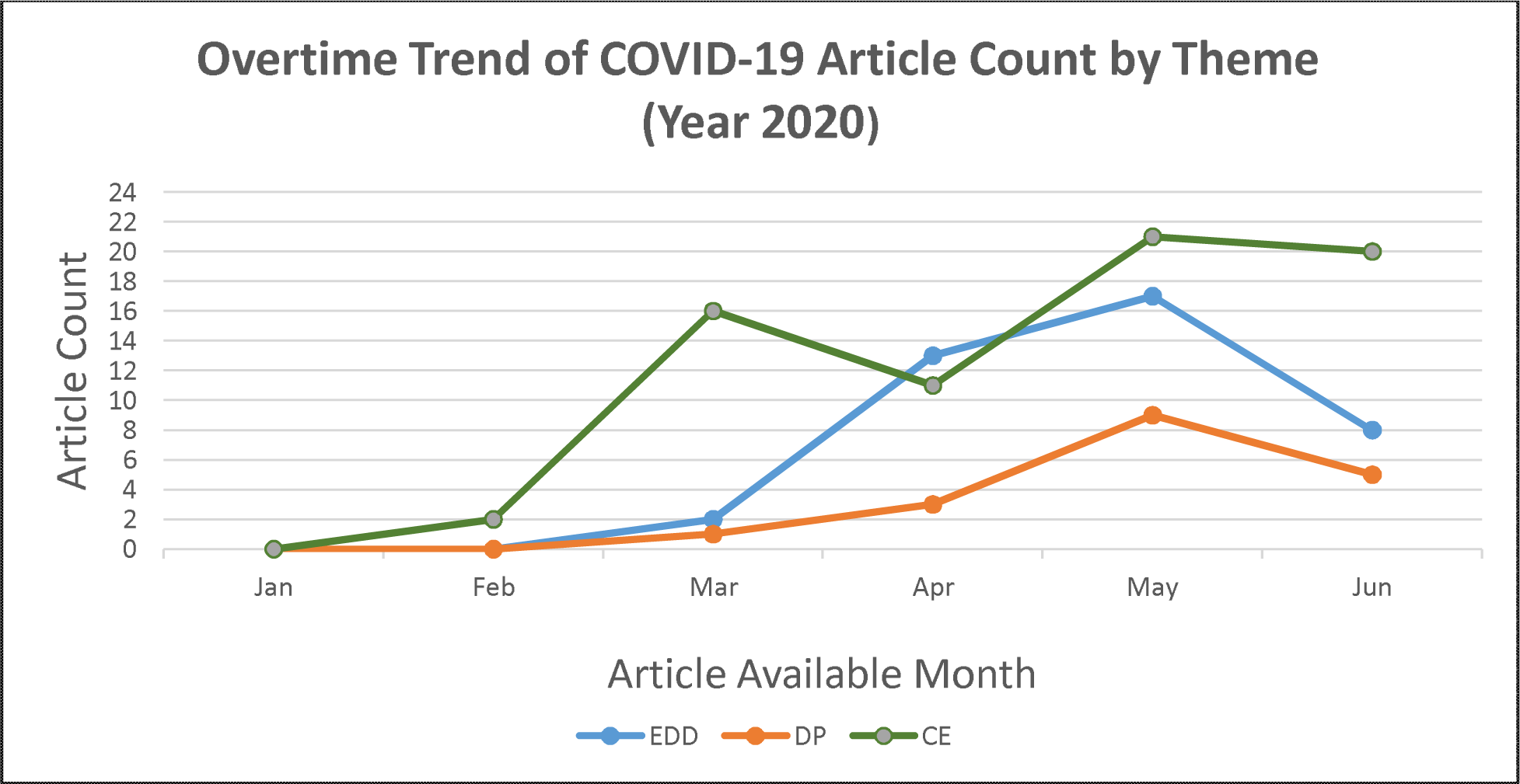
Overtime trend of COVID-19 articles that applied ML techniques made available online and categorized by the themes identified: 1) Computational Epidemiology (CE), 2) Early Detection and Diagnosis (EDD), and 3) Disease Progression (DP). For preprint articles, publication month of the latest version available as of query search date was used.

### Computational Epidemiology (CE)

The 70 studies that focused on epidemiological COVID-19 concerns were further classified into three categories 1) COVID-19 Disease Trajectory (CDT), 2) Molecular Analysis-Drug Discovery (MADD), and 3) Facilitate COVID-19 Response (FCR). These classifications were based on the aims of the study such as predicting outbreak, potential drug discoveries, policies, and other measures to stop the spread as shown in Table 2. Forty studies that focused on predicting COVID-19 peaks and sizes globally, and specific to a geographical location, estimating the impact of socioeconomic factors, and environmental conditions on the spread of the disease, and effectiveness of social distancing policies in containing the spread were grouped into the CDT. Twenty-two studies were grouped under the MADD based on the study approach in identifying existing drugs that have the potential to treat COVID-19, protein structure analysis, and predicting mutation rate in COVID-19 positive patients. The FCR includes 8 studies that emphasize on building tools to combat the ongoing pandemic such as building COVID-19 imaging repository, AI-enabled automatic cleaning and sanitizing tasks at healthcare facilities that might help clinical practitioners to provide timely services to the affected population. A majority of the studies in the CE theme used data either from social media (8 studies used data from Twitter, Weibo, or Facebook) or public data repositories such as NCBI, Drug Bank databases, and other health agencies data. Refer to Multimedia Appendix 2 for individual study details.

**Table 2.**
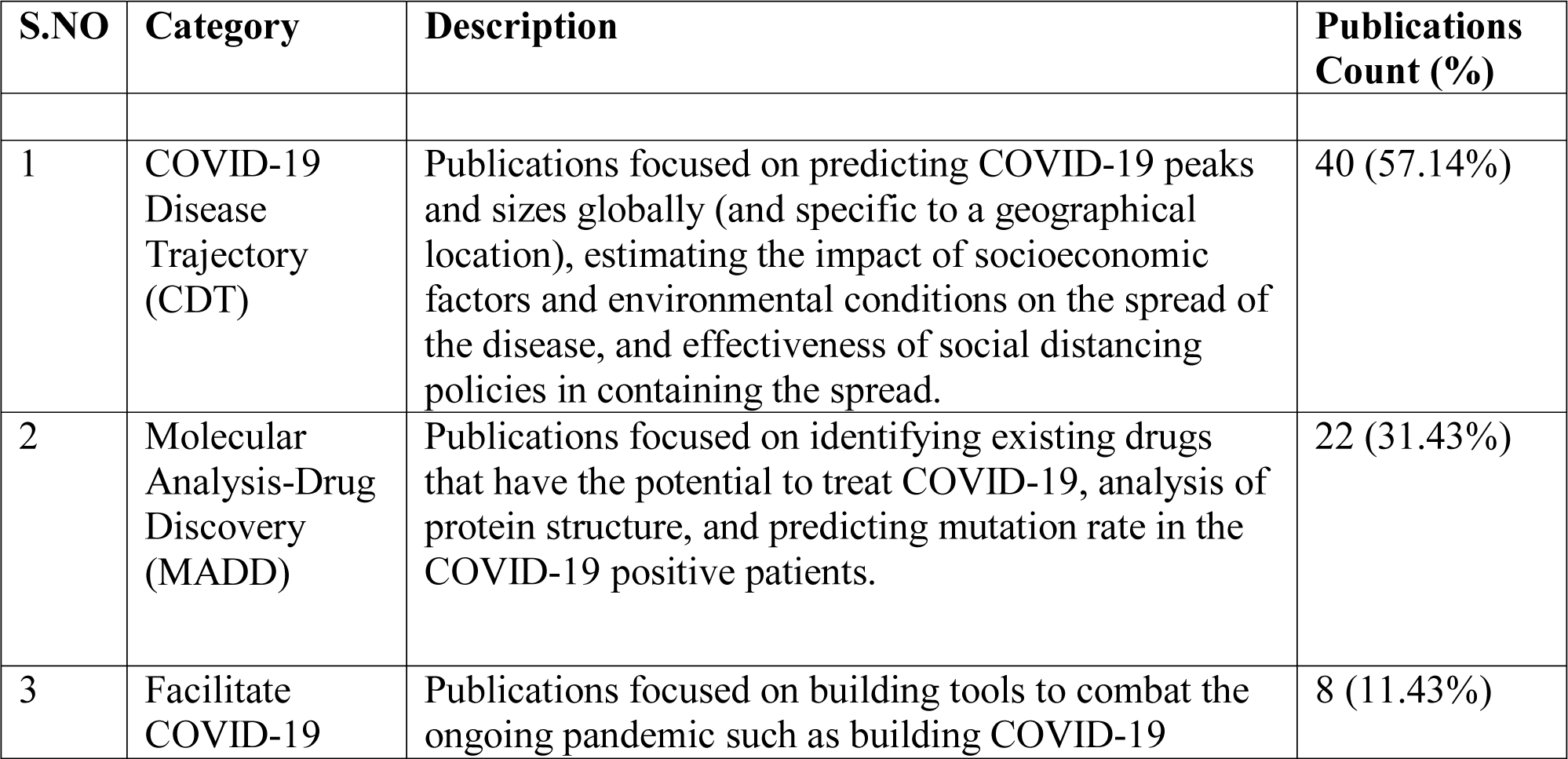
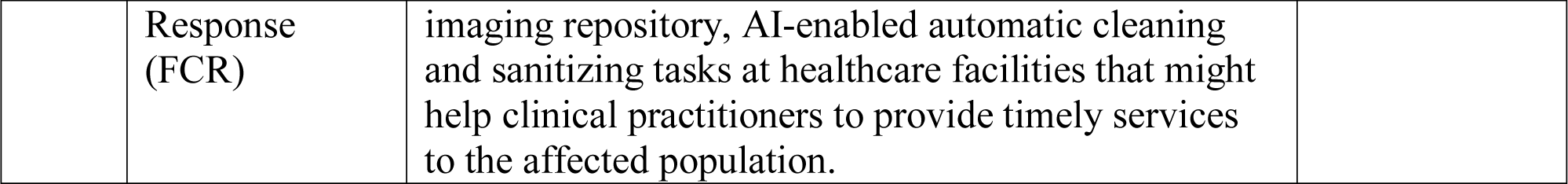
Computational Epidemiology (CE) publications classified into three categories 1) COVID-19 Disease Trajectory (CDT), 2) Molecular Analysis-Drug Discovery (MADD), and 3) Facilitate COVID-19 Response (FCR).

### Early Detection and Diagnosis (EDD)

We identified forty studies that primarily focused on diagnosing COVID-19 in patients with suspected infection mostly using chest radiological images such as Computed Tomography (CT), X-Radiation (X-Ray), and Lung Ultrasound (LUS). As shown in Table 3, twenty-three studies used X-Ray, fifteen used CT, one study used LUS, and finally, one study utilized non-imaging clinical data. Most studies used Deep Learning (DL) techniques to diagnose COVID-19 from radiological images. Nine studies employed ResNet, 4 studies used Xception, and 3 studies used VGG neural network models for either pre-training or as a diagnostic model. The study that used non-imaging clinical data to diagnose COVID-19 employed routine lab results captured in the Electronic Health Record (EHR) systems. Refer to Multimedia Appendix 3 for individual study details.

**Table 3.** Early Detection and Diagnosis (EDD) publications classified into four categories based on modality used for prediction: 1) X-Ray, 2) CT, 3) LUS, and 4) Non-Imaging Clinical data

| S.NO | Category | Description | Publications Count (%) |
| --- | --- | --- | --- |
| 1 | X-Ray | Publications focused on application of ML techniques to detect and differentiate COVID-19 positive patients from the general population using X-Ray images. | 23 (57.50%) |
| 2 | CT | Publications focused on application of ML techniques to detect and differentiate COVID-19 positive patients from the general population using CT images. | 15 (37.50%) |
| 3 | LUS | Publication focused on application of ML techniques to detect and differentiate COVID-19 positive patients from the general population using LUS images. | 1 (2.50%) |
| 4 | Non-Imaging Clinical data | Publication focused on application of ML techniques to detect and differentiate COVID-19 positive patients from the general population using non-imaging clinical data. | 1 (2.50%) |

### Disease Progression (DP)

We identified 18 studies that primarily focused on the prognosis of the disease in COVID-19 positive patients. We further classified these studies into 1) Risk Stratification: Publications focused on assessing the risk of disease progression (14 studies) and 2) Hospital Resource Management: Publications focused on predicting the need for hospital resources (4 studies) as shown in the Table 4. All studies in DP used demographic variables, 12 used comorbidities, and 11 used radiological images for analysis. Refer to Multimedia Appendix 4 for individual study details grouped under this theme.

**Table 4.** Disease Progression (DP) publications classified into two categories: 1) Risk Stratification and 2) Hospital Resource Management.

| S.NO | Category | Description | Publications Count (%) |
| --- | --- | --- | --- |
| 1 | Risk Stratification | Publications focused on assessing the risk of disease progression. | 14 (77.78%) |
| 2 | Hospital Resource Management | Publications focused on predicting the need for hospital resources. | 4 (22.22%) |

## Discussion

ML techniques will continue being used for monitoring, detection, and containment of the COVID-19 pandemic [55, 93, 129]. Our systematic review focused on 128 studies that applied ML methods and identified 3 themes: models developed to address issues central to Epidemiology; models that aid the diagnosis of patients with COVID-19; and models that help prognosis of COVID-19 patients.

### Computational Epidemiology (CE)

#### Molecular Analysis-Drug Discovery (MADD)

On average, the conventional drug discovery process takes 10 to 15 years with very low success rates [146]. Instead, drug re-purposing attempts have been made to explore similarities between the COVID-19 and other viruses such as Severe Acute Respiratory Syndrome (SARS) and Acquired Immunodeficiency Syndrome (AIDS) [147]. With the rapid accumulation of genetic and other biomedical data in recent years, ML techniques facilitate the analysis of already available drugs and chemical compounds to find new therapeutic indications [148].

The main protease (Mpro) of COVID-19 is a key enzyme in polyprotein processing, it plays an important role in mediating viral replication and transcription [149]. Several studies have applied ML techniques to identify drug leads that target the Mpro of severe acute respiratory syndrome coronavirus 2 (SARS-CoV-2), making it an attractive drug target [150, 151]. Ton et al [86] built a deep learning platform, Deep Docking (DD), which enables structure □ based virtual screening of billions of purchasable molecules in a short time. DD was applied to more than 1 billion compounds available from the ZINC15 library to identify the top 1,000 potential ligands for SARS CoV 2 Mpro protein. The proposed docking platform is a computationally cheaper and faster ML method compared to traditional docking methods, allowing faster screening of large chemical libraries, containing billions of compounds.

Beck et al [15] used a drug-target interaction ML model to identify top 10 commercially available drugs that could act on viral proteins of SARS-CoV-2. The ML model, Molecule Transformer-Drug Target Interaction (MT-DTI) used to predict binding affinity values between marketable antiviral drugs that could target COVID-19 proteins. The author claims, MT-DTI can accurately predict binding affinity based on chemical and amino acid sequences of a target protein without their structural information. Also, the study reveals that Atazanavir was the best chemical compound with K_d_ of 94.94 nM followed by Remdesivir (113.13 nM), Efavirenz (199.17 nM), Ritonavir (204.05 nM), and Dolutegravir (336.91 nM) against the SARS-CoV-2 3C-like proteinase. Computational drug repositioning ML models provide a fast, and cost-effective way to identify promising repositioning opportunities, and expedited approval procedures [148, 152].

Genome sequencing of various viruses is done to identify regions of similarity that may have consequences for functional, structural, or evolutionary relationships [153]. Due to heavy computational requirements of traditional alignment-based methods, alignment-free genome comparison methods are gaining popularity [153, 154]. A case study by Randhawa et al [83] proposed a ML based alignment-free approach for an ultra-fast, inexpensive, and taxonomic classification of whole COVID-19 virus genomes and can be used for classification of COVID-19 pathogens in real time.

#### Facilitate COVID-19 Response (FCR)

COVID-19 lockdown and home-confinement restrictions is having adverse impact on the mental well-being of the general population and specific to high-risk groups including health care workers, children, and older adults [155]. Several studies were done to understand and respond to these public health emergencies. Li et al [62] conducted a study using ML (Support Vector Machine) model and sentiment analysis to explore the impacts of COVID-19 on people’s mental health and to assist policymakers in developing actionable policies that could aid clinical practitioners. Weibo posts were collected before and after the declaration of the pandemic to build emotional score indicators and cognitive indicators. Key findings of the study reveal that after the declaration of COVID-19 in China, there has been a significant impact on increased negative emotions including anxiety and depression, increased sensitivity to social risks, and declined happiness and life satisfaction. Raamkumar et al [17] have used the Health Belief Model (HBM) [156] to determine public perception towards physical distancing posts from multiple public health authorities. The authors used DL (a variant of recurrent neural network) text classification model to classify Facebook comments related to the posts into four HBM constructs: perceived severity, perceived susceptibility, perceived barriers, and perceived benefits. Recent developments in the field of Natural Language Processing (NLP), BERT [157], XLNet [158], and other hybrid ML models have shown promising results in the field of sentimental analysis. Future studies should focus on these advanced techniques for improved social media content analysis.

There is a growing trend of collaborations among researchers and scientists around the world to combat the COVID-19 pandemic [159]. The emergence of COVID-19 encouraged public health agencies and the scientific community including journals and publishers to promote and ensure that research findings and data relevant to this outbreak are shared rapidly and openly [13, 160]. Peng et al [65] focused on creating a repository of COVID-19 chest X-Ray and CT images. The repository, COVID-19-CT-CXR, is publicly available that contains 1,327 CT and 263 X-Ray images (as of May 9, 2020) that are weakly labeled. The authors build a pipeline to automatically extract images from biomedical literature relevant to COVID-19 using a DL model. A recent effort by National Center for Advancing Translational Sciences (NCATS) to build a centralized national data repository on COVID-19 called National COVID Cohort Collaborative (N3C) is in progress [161]. The N3C will support collection and analysis of clinical, laboratory and diagnostic data from hospitals and health care plans. N3C along with the imaging repository such as the COVID-19-CT-CXR will accelerate clinical and translational research.

#### COVID-19 Disease Trajectory (CDT)

During the initial days of the COVID-19 spread, most research was focused on building mathematical models for estimating the transmission dynamics and prediction of COVID-19 developments [162, 163]. Specifically, Susceptible-Exposed-Infectious-Recovered (SEIR) and Auto-Regressive Integrated Moving Average (ARIMA) models and their extensions were widely adopted for projection of COVID-19 cases [164]. These models provided healthcare and government officials with optimal intervention strategies and control measures to combat the pandemic [164]. Yang et al [58] and Moftakhar et al [43] used ML models to fit statistical models SEIR and ARIMA respectively. Long-Short Term Memory (LSTM) model by Yang et al [58] and Artificial Neural Network (ANN) model by Moftakhar et al [43] had a good fit to the aforementioned mathematical models respectively. However, projections of both SEIR and ARIMA mathematical models had deviations less than ± 15% range of the reported data [164]. Future studies should try to fit ML techniques on both the SEIR and ARIMA models to reduce the projection error rate and to be prepared for second wave of COVID-19.

In early days of the pandemic, majority of the studies in our review predicted potential hotspots and outbreak trends using COVID-19 data from China expecting similar epidemic growth. However, the projections were off target due to varying containment policies enforced by different countries [164, 165]. Study by Yang et al [58] used ML technique to predict the COVID-19 epidemic peaks and sizes with respect to the containment polices. The study revealed that the continual enforcement of quarantine restrictions, early detection and subsequent isolation was most effective in the containment of the spread. Relaxing these policies would increase the spread by three-fold for a five-day delay in implementation and could cause a second peak. Such policies should be strictly enforced to prevent a second coronavirus outbreak.

#### Early Detection and Diagnosis (EDD)

Many countries ramped up the production of real-time reverse transcription polymerase chain reaction (RT-PCR) testing kits to diagnose COVID-19, and as of today, it remains the gold standard for confirmation [166]. However, the lab test suffers from low sensitivity as reported by several studies [166, 167]. Radiological images (CT and X-Ray) have been used by clinicians to confirm corona positive cases and it serves as an important complement to the RT-PCR test [168]. Several studies have reported that the use of chest CT for early-stage detection has proven to have a low rate of misdiagnosis and provide accurate results even in some asymptomatic cases [169].

Researchers started applying ML techniques on radiological scans to distinguish between the COVID-19 and non-COVID-19 cases, we identified 15 studies that used CT to detect COVID-19. One of the most cited studies Li et al [118], applied DL (COVNet) to differentiate COVID-19 and non-COVID-19 pneumonia CT scans. The Area Under the Receiver Operating Characteristics (AUROC) reported to identify COVID-19 on chest CT exam was 0.96 and to identify community-acquired pneumonia on chest CT exam was 0.95. The risks associated with using chest CT for diagnosis are 1) high radiation dose (7 mSv) compared to chest X-Ray (0.1 mSv) and 2) Chest CTs are more expensive compared to chest X-Ray [170, 171]. We identified 23 studies that used chest X-Ray and applied ML techniques to diagnose COVID-19 cases. A study by Apostolopoulos et al [89] applied a transfer learning strategy to train Convolutional Neural Network (CNN) models and then automated the detection of COVID-19 using chest X-Ray images. The model (VGG19) achieved an overall accuracy of 97.82% to detect COVID-19 on a dataset of 224 COVID-19, 700 pneumonia, and 504 normal X-Ray images. A similar study was done by Khan et al [115] using transfer learning and CNN (Xception) architecture with 71 layers that were trained on the ImageNet dataset. Their model (CoroNet) achieved an average accuracy of 87% to detect COVID-19 on a dataset of 284 COVID-19, 657 pneumonia (both viral and bacterial), and 310 normal chest X-Ray images. While chest X-Rays are cost-effective and have a much lower dose when compared to chest CTs, they are less sensitive, especially in the early stages of the infection, and also in respect of mild cases [172]. We recommend newer studies to develop ML models that can detect COVID-19 from the combination of CT and XRay images to aid clinical practitioners.

LUS have been proven useful during the 2009 influenza epidemic (H1N1) by accurately differentiating viral and bacterial pneumonia, and were found to have higher sensitivity to detect avian influenza (H7N9) when compared to chest X-Ray [173]. Though clinicians recommend the use of LUS imaging in the emergency room for diagnosis and management of COVID-19, its role is still unclear [174]. In our review, we identified one study by Roy et al [126] who used a deep learning model on annotated LUS COVID-19 dataset to predict disease severity. The results of the study were reported to be “satisfactory”.

In general, DL techniques are employed to improve prediction accuracy by training on large volumes of data [175]. In our review, several studies applied ML techniques, either using smaller imaging datasets specific to the organization, or mid-to-large dataset from publicly available repositories. However, there is a huge amount of cost associated with developing and maintaining such repositories [176]. To overcome the data size and cost limitations, Xu et al [108] proposed a decentralized AI architecture to build a generalizable ML model that is distributed and trained on in-house client datasets, eliminating the need for sharing sensitive clinical data. The proposed framework is in the early phase of adoption and needs technical improvements before it is widely employed by participating healthcare organizations.

An alternative to RT-PCR test, study by Joshi et al [113] proposed ML approach that utilize only Complete Blood Count (CBC) and gender to predict COVID-19 positivity. The authors build a logistic regression model on retrospective data collected from a single institute and validated on multi-institute data. Prediction of coronavirus positivity was reported to be C-statistic 78% and sensitivity 93%. The goal of the study was to develop a decision support tool that integrates readily available lab results from EHRs.

### Disease Progression (DP)

#### Hospital Resource Management

The novel coronavirus (COVID-19) pandemic has strained global healthcare systems, especially ICUs, due to hospitalized patients having higher ICU transfer rates [133]. Identification of hospitalized patients at high-risk in advance may help healthcare providers to plan and prepare for ICU resources (beds, ventilators, and staff, etc.) [177]. A study by Cheng et al [133] developed an ML-based model to predict ICU transfers within 24 hours of hospital admission. The Random Forest model was used for prediction and was based on variables: vital signs, nursing assessment, lab results, and electrocardiograms collected during the hospitalization. The overall AUROC of the model was reported to be 79.9%. Similar work was done by Shashikumar et al [139] to predict the need for ventilation in hospitalized patients 24 hours in advance. The prediction was not only limited to COVID-19 patients but also for generally hospitalized patients. The authors used 40 clinical variables: 6 demographic and 34 dynamic variables (including lab results, vital signs, Sequential Organ Failure Assessment (SOFA), comorbidity, and length of stay). In contrast to the traditional ML model used by Cheng et al [133], Shashikumar et al [139] resorted to a DL model (VentNet) for prediction with an Area Under the Curve (AUC) of 0.882 for the general ICU population and 0.918 for patients with COVID-19. Both the aforementioned studies relied on clinical variables for prediction whereas a study by Burian et al [130] combined clinical and imaging parameters for estimating the need for ICU treatment. The major finding of the study was, patients that needed ICU transfers had significantly increased Interleukin-6 (IL-6), C – reactive protein (CRP) and leukocyte counts and significantly decreased lymphocyte counts. All studies in this category applied ML techniques to facilitate the efficient use of clinical resources and help hospitals plan their flow of operations to fight the ongoing pandemic.

#### Risk Stratification

Developing a risk stratification mechanism among COVID-19 patients helps to facilitate timely assessments, allocate hospital resources, and appropriate decision making [178]. Jiang et al [136] model used demographics, vital signs, comorbidities, and lab results to predict patients that are likely to develop Acute Respiratory Distress Syndrome (ARDS). Of these variables, lab results alanine aminotransferase (ALT), the presence of myalgias, and elevated hemoglobin were the most predictive features. The overall accuracy of predicting ARDS was 80%. Moreover, using ALT alone, the model achieved an accuracy of 70%. However, the conclusion was drawn based on a limited patient (n = 53) set.

Yadaw et al [144] evaluated different ML models to classify COVID-19 patients into deceased or alive classes. The classification was based on five features: age, minimum oxygen saturation during the encounter, type of patient encounter, hydroxychloroquine use, and maximum body temperature. The study revealed age and minimum oxygen saturation during encounters were the most predictive features among different models. The author reports hydroxychloroquine as one of the top mortality predictors identified by XGBoost and Support Vector Machine (SVM) models; however, only a third of the patients in the dataset had been treated with this drug. Overall AUC was reported as 0.91. Ji et al [135] focused on early identification of COVID-19 positive patients that are likely to be at high-risk. Variables used for prediction were demographics, comorbidities, and lab results. The study identified a strong correlation between comorbidities and disease progression as supported by various other studies. The study further claims, a decrease in lymphocyte count and an increase in lactate dehydrogenase are related to disease progression. Overall AUC reported was 0.759. Both studies, Yadaw et al [144] and Ji et al [135] trained their respective ML models on the retrospective data and validated on prospective data.

Zhang et al [145] build a DL diagnostic and prognostic predictive model to detect COVID-19 and identified variables associated with risk factors for early intervention and monitoring. The study used 3,777 patients (5,468 CT scans) to differentiate coronavirus pneumonia from other pneumonia and normal controls. AUROC of the model was reported as 0.97. To improve image segmentation performance a DL model (DeepLabv3) was employed. The study further evaluated the effect of various drug treatments on lesion size and volume changes by comparing the differences between pre-treatment and post-treatment. The work claimed to provide a potential quantitative evaluation of the efficacy of drug treatment. The study employed five senior radiologists to annotate and review image segmentation, the availability of such resources is very expensive. To avoid the time-consuming annotation, Wang et al [143] used a transfer learning strategy to aid COVID-19 diagnostic and prognostic analysis. The study used a two-step transfer learning strategy, first, the model was trained on a large lung cancer CT dataset (4106 patient) along with Epidermal Growth Factor Receptor (EGFR) gene sequencing to learn associations between chest CT image and micro-level lung functional abnormality. Later, the model was trained and validated to differentiate COVID-19 from other pneumonia (AUC 0.87–0.88) and viral pneumonia (AUC 0.86). Moreover, the model classified patients into high-risk and low-risk categories to help manage medical resources and provide early interventions. For COVID-19 disease evaluation and risk stratification, Li et al [137] built a pulmonary disease severity score using X-Rays and neural network models. The score was computed as the Euclidean distance between the patient’s image and a pool of normal images using the Siamese neural network. The score predicted (AUROC 0.80) subsequent intubation or death within three days of hospital admission for patients that were initially not intubated.

This review has some inherent limitations. First, there is a possibility of studies missed due to the search methodology. Second, we removed five publications where full text was not available, and this may have introduced bias. Third, we included studies that were available as preprints. Finally, a comparison of ML model performance was not possible in the quantitative descriptive analysis as variables, sample size, and source of data was diverse across the studies. The current systematic review includes studies that were available online as of June 27^th^, as the pandemic progresses, we intend to write a second review on the studies published after the aforementioned date.

### Conclusions

In this systematic review, we assembled the current COVID-19 literature that utilized ML methods in the area of applications ranging from tracking, containing, and treating viral infection. Our study provide insights on the prospects of ML on the identified COVID-19 themes: epidemiology, early detection, and disease progression highlighting the important variables, data types, and available COVID-19 resources that can assist in facilitating clinical and translational research. Our study sheds light on the ML application as a potential drug discovery and risk stratification tool. The analysis of our review suggested that ML based diagnostic tools are highly accurate in detecting presence of the virus using radiological images and can be employed as a decision support tool.

## Data Availability

Data sharing/availability is not applicable to this article as it is a review article.

## Acknowledgments

This review did not receive any specific grant from funding agencies in the public, commercial, or not-for-profit sectors.

## Conflicts of Interest

None declared.

## Abbreviations

AI: Artificial Intelligence
AIDS: Acquired Immunodeficiency Syndrome
ALT: alanine aminotransferase
ANN: Artificial Neural Network
ARDS: Acute Respiratory Distress Syndrome
ARIMA: Auto-Regressive Integrated Moving Average
AUC: Area Under the Curve
AUROC: Area Under the Receiver Operating Characteristics
CBC: Complete Blood Count
CDT: COVID-19 Disease Trajectory
CE: Computational Epidemiology
CNN: Convolutional Neural Network
CRP: C – reactive protein
CT: Computed Tomography
CXR: Chest X-Ray
DD: Deep Docking
DL: Deep Learning
DP: Disease Progression
EDD: Early Detection and Diagnosis
EGFR: Epidermal Growth Factor Receptor
FCR: Facilitate COVID-19 Response
HBM: Health Belief Model
EHR: Electronic Health Record
ICU: Intensive Care Unit
IL-6: Interleukin-6
LOS: Length of Stay
LSTM: Long-Short Term Memory
LUS: Lung Ultrasound
MADD: Molecular Analysis-Drug Discovery
ML: Machine Learning
Mpro: Main Protease
MT-DTI: Molecule Transformer-Drug Target Interaction
N3C: National COVID Cohort Collaborative
NCATS: National Center for Advancing Translational Sciences
NCBI: National Center for Biotechnology Information
NLP: Natural Language Processing
PRISMA: Preferred Reporting Items for Systematic Reviews and Meta-analysis
RT-PCR: Reverse Transcription Polymerase Chain Reaction
SARS: Severe Acute Respiratory Syndrome
SARS-CoV-2: Severe Acute Respiratory Syndrome Coronavirus 2
SEIR: Specifically, Susceptible-Exposed-Infectious-Recovered
SOFA: Sequential Organ Failure Assessment
SVM: Support Vector Machine
XGBoost: Extreme Gradient Boosting
X-Ray: X-Radiation

## Multimedia Appendix 1

Query syntax for study search in all 3 databases (PubMed, CINAHL, and Web of Science).

## Multimedia Appendix 2

Details of 70 studies qualified under the Computational Epidemiology (CE) theme.

## Multimedia Appendix 3

Details of 40 studies qualified under the Early Detection and Diagnosis (EDD) theme.

## Multimedia Appendix 4

Details of 18 studies qualified under the Disease Progression (DP) theme.

## Notes

### Competing Interest Statement

The authors have declared no competing interest.

### Author Declarations

Not Applicable.Review of available evidence

